# COVID-19: Why SOLIDARITY and DisCoVeRy trials may fail to bring informative and timely results

**DOI:** 10.1101/2020.06.01.20118927

**Authors:** Mondher Toumi, Shuyao Liang, Monique Dabbous, Yitong Wang, Tingting Qiu, Ru Han

**Affiliations:** Public Health Department, Aix Marseille University, 27, bd Jean Moulin, 13385 Marseille Cedex 05, France

**Keywords:** SARS-CoV-2, COVID-19, SOLIDARITY trial, DisCoVeRy trial

## Abstract

**Objective:** The SOLIDARITY and DisCoVeRy trials were launched to facilitate the rapid worldwide comparison of the efficacy and safety of treatments against COVID-19. This study aimed to review the trial designs of SOLIDARITY and DisCoVeRy and their feasibility to generate high-quality evidence.

**Method:** A systematic search of the European Clinical trial registry, the U.S. National Library of Medicine ClinicalTrials.gov, and the World Health Organization’s (WHO) International Clinical Trials Registry Platform (ICTRP) was conducted on May 10^th^, 2020 to identify the study details of the SOLIDARITY and DisCoVeRy trials. A supplementary search of PubMed, WHO’s website, French authorities’ websites, and Google search engine was conducted. A critical review was performed on the findings.

**Results:** The DisCoVeRy trial design was detailed consistently in both the European and the US clinical rial registries. SOLIDARITY was registered on ICTRP, with country-specific information reported on country-level registry platforms. The DisCoVeRy trial’s design appears to be ideal from the methodological perspective. Both trials appear difficult to implement, impractical, and disconnected from the pandemic reality. This is consistent with the apparent failure of the trials to deliver conclusions before the end of the pandemic.

**Conclusion:** Both trials constitute an interesting initiative yet may lack the resources to support a high-quality implementation. The authors call for a pandemic task force, with various experts on the front-line of COVID-19, to inform policy-makers to make effective decisions that may not be based on traditional, methodological state-of-the-art evidence, but rather pragmatic and revisable decisions reflecting emerging evidence for the benefit of patients and society.

## 1. Introduction

A new pneumonia epidemic was initially reported in China in December of 2019. The discovered virus and the associated disease were named Severe Acute Respiratory Syndrome Coronavirus 2 (SARS-CoV-2) and Coronavirus Disease 2019 (COVID-19), respectively. On March 12^th^, 2020, the World Health Organization (WHO) declared COVID-19 as a pandemic due to its fast-worldwide spread. The infection rate as well as the mortality rate remain unknown at the time of this publication. However, various studies suggest the infection rate to be between 10% [1] and 25% [2], while the mortality rate may range between 0.5% and 10% [3]. As of May 17^th^,2020, the global mortality rate is estimated to be 6.78% [4], which is likely to be an over-estimation as the denominator remains unknown, presently.

No vaccine is available and no drug with proven efficacy has been approved. Only in-vitro evidence of proposed treatments has shown potential antiviral activity against SARS-CoV-2. However, in-vitro evidence is not predictive of clinical efficacy for virus infections. Robust clinical evidence on efficacy and safety is needed for the benefit-risk assessment in the approval of potential treatments for COVID-19. On March 18^th^, 2020, the WHO Director-General announced the launch of a multinational Phase III-IV clinical trial called SOLIDARITY to facilitate the rapid worldwide comparison of unproven treatments. Four existing antiviral and anti-inflammatory agents are to be compared with standard-of-care (SoC) in the SOLIDARITY trial to evaluate their relative effectiveness and safety against COVID-19 through the enrollment of hospitalized patients in multiple countries. Other treatment candidates can be adaptive and added into the trial with emergence of new evidence. As a part of SOLIDARITY trial program, France launched a satellite European trial called DisCoVeRy, which intends to analyze the efficacy and safety of treatment options for patients within a limited time frame. The DisCoVeRy study was financed by the Ministries of Higher Education, Research, and Innovation (MESRI) and of Health and Solidarity (MSS), and was coordinated by the French National Institute of Health and Medical Research (Inserm) as part of the REACTting consortium. The list of potential drugs for the trial is based on the subset of experimental treatments classified as top priority by the WHO. This study aimed to review the trial designs of SOLIDARITY and DisCoVeRy, analyze their strengths and weaknesses, and the feasibility of the two trials.

## 2. Method

A systematic search of the European Clinical trial registry, the U.S. National Library of Medicine ClinicalTrials.gov, and the WHO’s International Clinical Trials Registry Platform (ICTRP) was conducted on May 10^th^, 2020 to identify the study designs of the SOLIDARITY and DisCoVeRy trials. With regard to the SOLIDARITY study, the trials reported at national level in clinical trial registries were also identified. A supplementary search of PubMed, WHO’s website, French authorities’ websites including MESRI, MSS and Inserm, and Google search engine using the keywords of “SOLIDARITY trial” and “DisCoVeRy trial” was conducted to identify additional information on the progress of the two trials.

## 3. Results

The different constituents of the study design of the SOLIDARITY and DisCoVeRy trials in clinical trial registries were reported (Table 1). The nation-specific data on the study design of the DisCoVeRy trial was further reported (Table 2). Considering the adaptive design of the SOLIDARITY and DisCoVeRy trials, the up-to-date parameters modified in accordance with observations in the trials could be followed up in the registries mentioned in this study.

**Table 1.** Comparison of study design between Solidarity and Discovery trial.

|  | Solidarity trial | Discovery trial |
| --- | --- | --- |
| Trial ID | PER-010-20 | NCT04315948 |
| Trial title | Solidarity: An International Randomized Controlled Trial to Evaluate Non-Licensed Covid-19 Treatments in Addition to Standard of Care among Hospitalized Patients | Multi-centre, Adaptive, Randomized Trial of the Safety and Efficacy of Treatments of COVID-19 in Hospitalized Adults (DisCoVeRy) |
| Planned date of enrolment | 03/04/2020 | 22/03/2020 |
| Recruitment status | Not Recruiting | Recruiting |
| Date of last update | 20/04/2020 | 29/04/2020 |
| Target size | 50,000 | 3,100 |
| Planned countries | India; Iran; Thailand; Spain; Norway; Switzerland; South Africa; Argentina; Peru; Bahrain; Norway; Finland; Canada | Belgium, France, Germany Luxembourg, the Netherlands, Spain, Sweden, and the United Kingdom |
| Study type | Adaptive RCT | Adaptive RCT |
| Masking | Open Label | Open Label |
| Intervention model | Parallel assignment in Norway and Canada; no parallel and no cross over group in Spain, Italy, Norway and Finland | Parallel assignment |
| Phase | Phase 3/4 | Phase 3 |
| Inclusion Criteria | 1. $\geq 18$ years of age<br>2. Has laboratory-confirmed SARS-CoV-2 infection as determined by PCR, or other commercial or public health assay in any specimen prior to randomization.<br>3. Hospitalized at a participating centre | 1. Adult $\geq 18$ years of age at time of enrolment.<br>2. Has laboratory-confirmed SARS-CoV-2 infection as determined by PCR, or other commercial or public health assay in any specimen $< 72$ hours prior to randomization.<br>3. Hospitalized patients with illness of any duration, and at least one of the following:<br>Clinical assessment (evidence of rales/crackles on exam) AND SpO2 $\leq 94\%$ on room air, OR<br>4. Acute respiratory failure requiring mechanical ventilation and/or supplemental oxygen.<br>Women of childbearing potential must agree to use contraception for the duration of the study. |
| Exclusion Criteria | 1. Severe co-morbidity with life expectancy $< 3$ months according to investigators assessment<br>2. Known intolerance to the available study drugs<br>3. Pregnancy or breast feeding | 1. Refusal to participate expressed by patient or legally authorized representative if they are present<br>2. Spontaneous blood ALT/AST levels $> 5$ times the upper limit of normal.<br>3. Stage 4 severe chronic kidney disease or requiring dialysis (i.e. eGFR $< 30$ mL/min)<br>4. Pregnancy or breast-feeding.<br>5. Anticipated transfer to another hospital, which is not a study site within 72 hours.<br>6. Patients previously treated with one of the antivirals evaluated in |
|  |  | the trial (i.e. remdesivir, interferon β-1a, lopinavir/ritonavir, hydroxychloroquine) in the past 29 days<br>7. Contraindication to any study medication including allergy<br>8. Use of medications that are contraindicated with lopinavir/ritonavir i.e. drugs whose metabolism is highly dependent on the isoform CYP3A with narrow therapeutic range (e.g. amiodarone, colchicine, simvastatine).<br>9. Use of medications that are contraindicated with hydroxychloroquine: citalopram, escitalopram, hydroxyzine, domperidone, pipéraquine.<br>10. Human immunodeficiency virus infection under highly active antiretroviral therapy (HAART).<br>History of severe depression or attempted suicide or current suicidal ideation |
| Intervention (dosing, if available) | 1. Remdesivir<br>2. Lopinavir/ritonavir (orally twice a day for 14 days)<br>3. Lopinavir/ritonavir (orally twice a day for 14 days) and Interferon Beta-1A (daily injection for 6 days)<br>4. Hydroxychloroquine (oral loading dose, then 200 mg orally twice daily for 10 days) | 1. Remdesivir (200 mg on Day 1, followed by a 100 mg once daily for the duration of the hospitalization up to 10 days total course)<br>2. Lopinavir/ritonavir (400 lopinavir mg/100 mg ritonavir every 12 h for 14 days in tablet or administered as a 5-ml suspension every 12 h for 14 days via a pre-existing or newly placed nasogastric tube)<br>3. Lopinavir/ritonavir (400 lopinavir mg/100 mg ritonavir every 12 h for 14 days in tablet or administered as a 5-ml suspension every 12 h for 14 days via a pre-existing or newly placed nasogastric tube) and Interferon Beta-1A (44 μg for a total of 3 doses in 6 days)<br>4. Hydroxychloroquine (400 mg twice daily for one day followed by 400 mg once daily for 9 days) |
| Control (dosing, if available) | Standard of Care | Standard of Care |
| Primary outcome | All-cause mortality at discharge | Percentage of subjects reporting each severity rating on a 7-point ordinal scale in 15 days |
| Responsible party | The United Nations Foundation, the Swiss Philanthropy Foundation, and the World Health Organization (WHO) | Institut National de la Santé Et de la Recherche Médicale, France |

*RCT: randomized controlled trial*

### 3.1. DisCoVeRy trial description

The study design of the DisCoVeRy study was reported consistently and elaborately in both the U.S. National Library of Medicine ClinicalTrials.gov registry [5] and the EU Clinical Trials Register [6].

#### 3.1.1. Study objective

The objective of the DisCoVeRy trial is to assess the efficacy and safety of 4 interventions against the SoC in COVID-19 for hospitalized adult patients.

#### 3.1.2. Timelines

The study start date was reported to be the 22^nd^ of March 2020, with the study completion estimated to be March 2023.

#### 3.1.3. Study design

DisCoVeRy is a European adaptive, randomized, multicentric open 5 parallel arms trial. Randomization is balanced as 1:1:1:1:1. The target sample size is 3100 patients enrolled, with 620 patients per treatment arm.

Patients are stratified according to geography and to the level of severity (moderate or severe) according to well-predefined criteria.

It is not clear what elements of the adaptive design will be considered for adjustment of the trial design by the study monitoring committee. Based on the available information on the U.S. National Library of Medicine’s ClinicalTrial.gov database, the adjustments are likely related to interventions that may be revised based on milestone results predefined in the adaptive study design. The number of patients and population do not seem to be considered as part of the changes to occur in the adaptive design.

The study duration per patients is not specified. However, the last end point is measured at day 90, suggesting it is the study end for each individual patient.

The study si being conducted in 34 centers, and patients are being recruited from Belgium, France, Germany, Luxembourg, Netherlands, Spain, Sweden, and the United Kingdom (UK).

#### 3.1.4. Intervention

Four interventions including Remdesivir, the combination of Lopinavir and Ritonavir, the combination of Lopinavir, Ritonavir and Interferon Beta, and Hydroxychloroquine, combined with or without SoC are compared to SoC alone. The dosages are well-specified while the duration of treatment may differ between patients for certain products. The SoC is not clearly defined and is adjusted based on country-specific protocol.

#### 3.1.5. Population

The population of the DisCoVeRy trial includes hospital-admiitted patients with Covid-19 and hypoxemia based on clinical assessment and laboratory testing, and with or without respiratory failure.

#### 3.1.6. Primary endpoints

In the DisCoVeRy trial, the primary endpoint reported is severity rating at day 15 on a 7-point ordinal scale:

- Not hospitalized, no limitations on activities;
- Not hospitalized, limitation on activities;
- Hospitalized, not requiring supplemental oxygen;
- Hospitalized, requiring supplemental oxygen;
- Hospitalized, on non-invasive ventilation or high flow oxygen devices;
- Hospitalized, on invasive mechanical ventilation or Extracorporel membrane oxygenation (ECMO);
- Death.

#### 3.1.7. Secondary endpoints

Twenty-three secondary endpoints are reported for the DisCoVeRy trial.

The first secondary endpoint is the same severity ranking as primary endpoint, but is assessed 6 times at day 3, 5, 8, 11, 15 and 29, respectively. The first secondary endpoint is complemented by 3 sub-endpoints including time to improve, subject clinical status and mean change in the severity ranking, of which, two are assessed 5 and 6 times, respectively. This already accounts for 48 secondary measures for the first secondary endpoint.

The second secondary endpoint is the time to discharge or to a NEWS of ≤ 2, depending on which occurs first and is measured 6 times at day 3, 5, 8, 11, 15 and 29, respectively.

When considering sub-endpoints and different timeframes, in total there are 76 secondary end points. Five additional endpoints are listed, of which, 2 are measured 6 separate times, and 4 are measured 4 separate times, resulting in an additional 28 endpoints. Therefore, the total number of endpoints collected is 104 when different timeframes are counted.

#### 3.1.8. Sample size calculation and statistics

No information was available on sample size calculation and statistics. Therefore, it is unclear how statistical analysis will be performed, especially when no statistical analysis is proposed for the secondary endpoints.

### 3.2. SOLIDARITY trial description

The study design of the SOLIDARITY trial was reported on the WHO’s ICTRP [7].

The SOLIDARITY and DisCoVeRy trials mostly share the same characteristics, such as study design, interventions, and population selection. The SOLIDARITY trial allowed for country-specific study customization, which may have significantly deviated from the original protocol (Table 2).

**Table 2.**
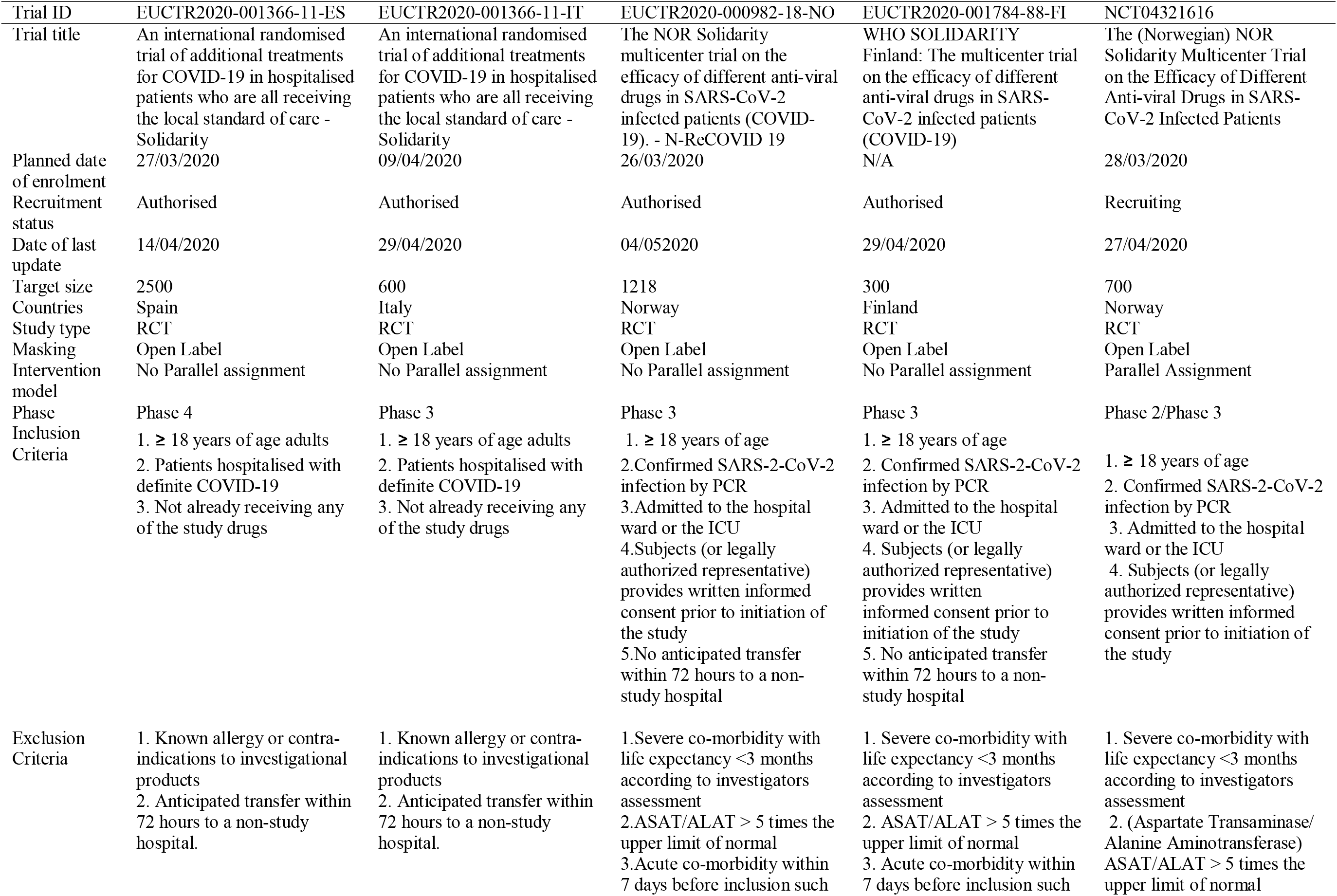
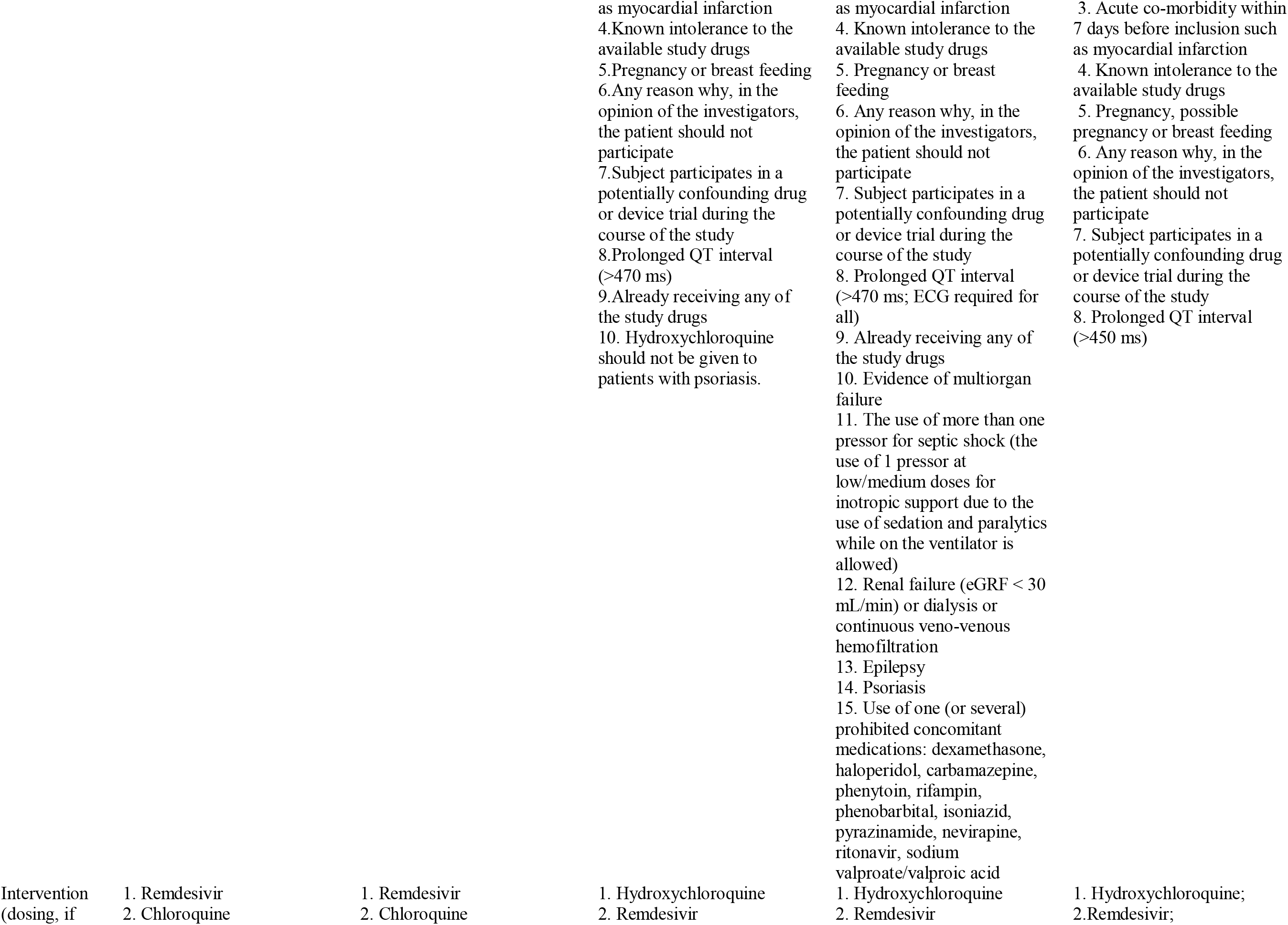
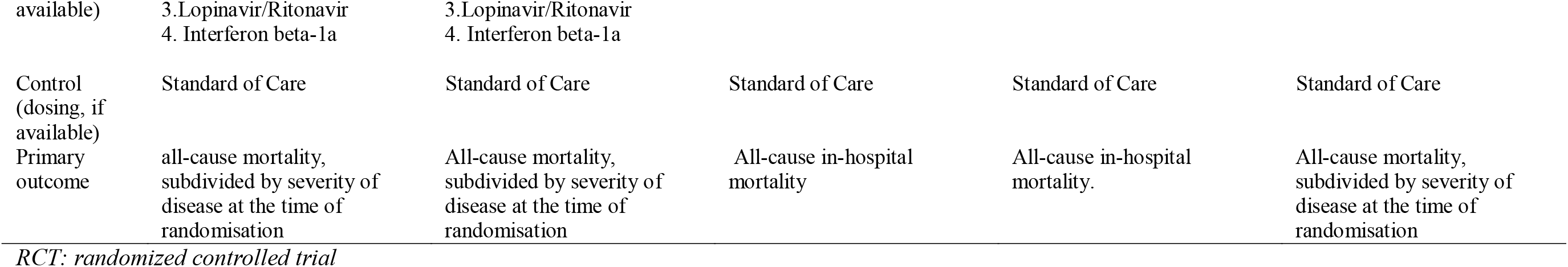
Solidarity trial conducted in different countries.

The SOLIDARITY trial’s population size is reported to be 50,000 patients, while it is reported to be 3,100 for the DisCoVeRy trial.

Primary endpoints vary significantly between the SOLIDARITY trial and the DisCoVeRy trial as the primary endpoint for the SOLIDARITY trial is all-cause mortality, while the primary endpoint for the DisCoVeRy trial remains severity rating at day 15 on a 7-point ordinal scale.

National customization has resulted in variations in secondary endpoints considered in the trial design and implementation of SOLIDARITY. The Canadian customization of the SOLIDARITY trial has considered a broad range of secondary endpoints more similar to those considered in France where severity ranking assessed with subject clinical status is being evaluated, which were not considered in Italy and Spain. It is unclear how these studies will be pooled and used to reach a final consistent conclusion.

With the exception of these points of differentiation, very little has been reported on the SOLIDARITY trial that could inform on further discrepancies.

## 4. Discussion

### 4.1. DisCoVeRy trial

It is evident that the DisCoVeRy trial was carefully designed from a theoretical perspective and if it would have been double-blinded it would be considered as a state-of-the-art trial. However, the theoretical perspective seems to have been disconnected from the reality of the pandemic.

Indeed, when healthcare systems are overloaded during an outbreak, it may be impossible to run a trial without a strong commitment from healthcare professionals (HCP) paying a high cost on their private time as they are exhausted and overloaded. Healthcare systems may have neither the resources nor the budget to implement such trials. Furthermore, the time dedicated to these trials may be at the patients’ expense, while physicians’ primary goal is to treat patients not to conduct trials. Even though trials are critical to HCPs, their priority will remain to treat patients queuing at the hospital gate.

The very large number of endpoints, more than 100, may be a strong disincentive to participate in the DisCoVeRy trial when HCPs may be unable to address the minimal requirements. A simple trial with mortality and time to clear virus load endpoints may gain more participation from HCPs.

The randomization may have also created a challenging situation for enrolled patients. Patients in a situation presented by the media as dramatic in outcomes may not easily accept to be part of a trial where they may be allocated to no active therapy while 4 potential effective treatments are tested, with most of them licensed and accessible without being part of the trial. The randomization between active arms and SoC may have been discouraging patients to participate to the trial. It has been reported by several investigators that patients wanted to access medicine and not to participate in a trial. Moreover, the large publicity around the likely ineffectiveness and potential adverse events of hydroxychloroquine (HCQ) made a randomized trial unacceptable for patients [8].

A study with an external arm not randomized for the SoC may have gained more traction with patients and HCPs. In such a case, a propensity-matched comparison could have been considered and been very informative in supporting decision-making in the light of a large sample size expected for such a study. It may not appear to meet generally accepted standards from the methodological perspective, but in order for the requested effect size to be relevant in the context of a pandemic, such a trial design could have been appropriate. A similar study conducted by Geleris et al to estimate the association between HCQ treatment and intubation or death rate of hospitalized patients with COVID-19 has been published. The propensity-score methods were used to reduce the effects of confounding in the baseline information of patients. A multivariable regression model with inverse probability weighting according to the propensity score was used and showed there was no significant association between HCQ use and the risk of intubation or death. A series of analyses using several propensity-score approaches were also performed to confirm the results and that the findings were consistent. Therefore, proven conclusions could be drawn from such a study design considering propensity-matched comparison [9].

Alternatively, a cluster randomization between centers willing to participate in the study and centers not willing to expose patients to unproven therapies could have maintained randomization on the center level, rather than the patient level. Again, such an approach may appear to be imperfect and not up to generally accepted standards, yet it could have very well addressed such an issue arising from the allocation of patients to unproven therapies.

Patients with well-established COVID-19 and hypoxemia may be, in most cases, exposed to the risk of a cytokine storm [10, 11], for which all the antiviral treatments currently experimentally used will not be effective for severe inflammatory injuries, with the exception of the interferon Beta (IFN-β) arm.

However, IFN-β may not be the most effective when other strong anti-cytokines storm exist, and others have been approved for CAR-T Cell-related cytokine storms, such as Tocilizumab [12]. The latest version of Chinese national guidelines for the treatment of COVID-19 indicates a 93% SaO_2_ threshold to prescribe anti-cytokine release syndrome therapies when considered in combination with anti-viral therapies [13].

Therefore, this population is too advanced to benefit from antiviral therapies and may benefit more from anti-cytokine release syndrome drugs, such as tocilizumab, rituximab or dupilumab. However, the protocol of the DisCoVeRy trial did not plan a priori that the population in the trial may be adjusted, therefore, an amendment would be required for such population changes and may not be compliant with good clinical practice (GCP). It could be planned a priori, rendering the trial compliant with GCP.

In relation to the interventions selected for the trial, debates flourished among experts suggesting that HCQ was not part of the initial protocol while at that time it was the only intervention with evidence, even though the evidence was weak. This was later revised and HCQ was finally, after intensive debates, included in the trial as one of the intervention tested [14, 15].

The time of the expected final trial results, 3 years from trial initiation, in 2023, may have discouraged many potential investigators. In the context of a pandemic, presenting a state of emergency and large number of patients, HCPs needed a fast and simple study to assess if any of the tested interventions were effective. A 3 year wait period for results seems to be totally disconnected from the sense of urgency among all practitioners, the public, and decision-makers.

The difference in healthcare systems and the levels of HCP overload was not considered when designing the trial. Implementation of such a trial may be too complex and burdensome in overloaded healthcare systems where HCPs are experiencing burnouts, complications and/or even death associated with COVID-19.

Ultimately, the likelihood that several European countries will adhere to a trial with French ownership and which was widely-communicated as the solution to such a large problem may represent a political issue, resulting in the hesitance of other countries to be engaged in such a project. It may seem difficult to Germany and the UK, for example, to accept that France has claimed to initially have paved the way for a solution to studying treatments for the COVID-19 pandemic, while potentially appearing to be followers and contributors. If the protocol had been developed jointly, this may have resulted in a delay for establishing and implementing such a study, however, it may have resulted in enhanced cooperation and adoption.

### 4.2. SOLIDARITY trial

The SOLIDARITY trial provides a very high-level design and allows for individual countries willing to participate in the study the ability to customize the protocol.

Customization has resulted in high heterogeneity of adoption and implementation, rendering it difficult to understand how to pool the data and interpret results.

Background therapies, such as SoC, may be different and the variation in available resources to run such a trial may well translate to a difference in quality of adaptation and implementation.

The sample size of 50,000 patients resembles more of an all-inclusive strategy, rather than a conscientiously scientific sample power calculation. Indeed, a lot of attention was gained relating to the massive, all-inclusive characteristic of this trial, rather than on the purpose and the actual, expected outcomes [16–18].

Each country customized the protocol and considered even more patients than the whole study, itself. For example, Italy and Spain anticipated to recruit 600 and 2500 patients, respectively. These numbers seem random and not supported by the reality of the situation in both countries. Countries, such as Finland and Norway, pragmatically customized the estimated patient numbers of the trial.

### 4.3. Current progress and next steps

There is little information on how the SOLIDARITY trail is progressing, with no information released on the quality of the evidence collected.

More information is available on the DisCoVeRy trial. However, this should be interpreted with caution. Only 25% of patients have been enrolled, while the epidemic is slowing down. No European countries have joined the project, except for Luxembourg with 1 patient as of 4^th^ May 4^th^, 2020 [19]. The UK has launched its own study called Recovery [20], considered by the British experts as the largest study yet on COVID-19 [21].

It is unclear what will result from the DisCoVeRy trial in 3 years when several more pragmatic studies may have already shown results supporting that HCQ does not work in hospitalized COVID-19 patients, especially severe patients [9, 22]. Remdesivir has gained marketing authorization for emergency use from the FDA in the US [23] based on one placebo-controlled randomized clinical trial and one open-label trial, and several more ongoing agile studies [24–28].

At the time of the results, an effective vaccine may well be available according to the FDA [29, 30] and EMA in early 2021 [31, 32]. The estimated completion date for the DisCoVeRy trial, March 2023, appears to be too late.

## 5. Conclusion

With the SOLIDARITY trial, WHO launched an interesting initiative, yet may have been lacking the resources to secure aligned, high-quality implementation.

France launched a European branch of the SOLIDARITY trial called DisCoVeRy. It may be argued that the study lacks pragmatism and is too complex. The DisCoVeRy trial ignores patients’ behavior in a pandemic with very high-perceived fatality risk. Therefore, patients tend to resist entering a trial with a placebo-like arm.

The very intense workload of clinicians requires that they manage their time critically, dedicating all their time to treat patients, rather than to conduct trials at the expense of patient management.

The long window to finalize this study may also have been a disincentive. From now until 2023, there will have been several telling trials already made available. It is very likely that a vaccine may have even reached the market by March 2023. Indeed, 3 additional SARS-Coronavirus-19 circulations may be experienced before the study results are available.

Finally, it may have been preemptive of French scientists and French health authorities to expect a very close collaboration under French leadership, while all EU Member States, including France, have handled this pandemic from unilateralistic perspectives.

In the context of a pandemic, well-established experts, methodologists, and the health authorities’ administrations traditionally operating on long term visions may not be the entities or pathways to launch agile, pragmatic, and effective research. There is a need to have such entities, authorities, and resources experienced in such catastrophic situations to be able to inform the policy decision-makers and take effective decisions that may not meet traditionally and generally accept standards from methodological perspective, but which may ultimately work for the benefit of patients and the society. Ultimately, this is a call for a pandemic task force with various experts from those handling the day-to-day research and administrative tasks to engage with such research in the context of the COVID-19 pandemic.

## Data Availability

A systematic search of the European Clinical trial registry, the U.S. National Library of Medicine ClinicalTrials.gov, and the International Clinical Trials Registry Platform (ICTRP) in WHO was conducted on May 10th, 2020 to identify the study designs of the SOLIDARITY and DisCoVeRy trials. With regard to the SOLIDARITY study, the trials reported at national level in clinical trial registries were also identified. A supplementary search of PubMed, the website of WHO and French authorities including MESRI, MSS and Inserm, and Google search engine using the keywords of SOLIDARITY trial and DisCoVeRy trial was conducted to identify additional information on the progress of the two trials.

## Declarations

### Funding

This study did not receive any specific grant from funding agencies in the public, commercial, or not-for-profit sectors.

#### Ethical Approval

Not required as this study used clinical trial registry platforms [European Clinical trial registry, the U.S. National Library of Medicine ClinicalTrials.gov, and the WHO’s International Clinical Trials Registry Platform (ICTRP)], PubMed, WHO website, French authorities’ websites including MESRI, MSS and Inserm, and Google search engine.

#### Authors’ contributions

Mondher Toumi contributed to the study conception, design, analysis and interpretation of data and original draft preparation. Shuyao LIANG contributed to the study conception, acquisition of data, and original draft preparation. Monique DABBOUS contributed to editing and review of the article. Yitong WANG, Tingting QIU and Ru HAN contributed to the study conception, acquisition of data and review of the article. All authors read and approved the final manuscript.

## References

1. Streeck, H., et al., Vorläufiges Ergebnis und Schlussfolgerungen der COVID-19 Case-Cluster-Study (Gemeinde Gangelt). Preprint published online on, 2020: p. 04–09.

2. Stedman, M., et al., A phased approach to unlocking during the COVID 19 pandemic–Lessons from trend analysis. International Journal of Clinical Practice, 2020: p. e13528.

3. Johns Hopkins Coronavirus Resource Center. How does mortality differ across countries? 2020 [cited 2020 May 18th]; Available from: https://coronavirus.jhu.edu/data/mortality.

4. World Health Organization (WHO). WHO Coronavirus Disease (COVID-19) Dashboard. [cited 2020 May 17th]; Available from: https://covid19.who.int/?gclid=CjwKCAjwwYP2BRBGEiwAkoBpAqWJDbCXHuIn8JbhNvGrZHXoaUWwmdclCgtdeOWPgzYsxjZ4sVylRoCKgkQAvD_BwE.

5. ClinicalTrials.gov. Trial of Treatments for COVID-19 in Hospitalized Adults (DisCoVeRy). 18 May 2020]; Available from: https://clinicaltrials.gov/ct2/show/NCT04315948.

6. EU Clinical Trials Register. Multi-centre, adaptive, randomized trial of the safety and efficacy of treatments of COVID-19 in hospitalized adults. 18 May 2020]; Available from: https://www.clinicaltrialsregister.eu/ctr-search/trial/2020-000936-23/FR.

7. International Clinical Trials Registry Platform (ICTRP). SOLIDARITY: AN INTERNATIONAL RANDOMIZED CONTROLLED TRIAL TO EVALUATE NON-LICENSED COVID-19 TREATMENTS IN ADDITION TO STANDARD OF CARE AMONG HOSPITALIZED PATIENTS. 18 May 2020]; Available from: https://www.ins.gob.pe/ensayosclinicos/rpec/recuperarECPBNuevoEN.asp?numec=010-20.

8. Sciama, Y. Is France’s president fueling the hype over an unproven coronavirus treatment? [cited 18 May 2020; Available from: https://www.sciencemag.org/news/2020/04/france-spresident-fueling-hype-over-unproven-coronavirus-treatment.

9. Geleris, J., et al., Observational Study of Hydroxychloroquine in Hospitalized Patients with Covid-19. New England Journal of Medicine, 2020.

10. Schett, G., M. Sticherling, and M.F. Neurath, COVID-19: risk for cytokine targeting in chronic inflammatory diseases? Nature Reviews Immunology, 2020. 20(5): p. 271–272.

11. Toumi, M. and S. Aballea, Commentary on “Hydroxychloroquine and azithromycin as a treatment of COVID-19: results of an open label non-randomized clinical trial” by Gautret et al. Journal of Market Access & Health Policy, 2020. 8(1): p. 1758390.

12. Le, R.Q., et al., FDA approval summary: tocilizumab for treatment of chimeric antigen receptor T cell induced severe or life threatening cytokine release syndrome. The oncologist, 2018. 23(8): p. 943.

13. National Health Commission & State Administration of Traditional Chinese Medicine. Diagnosis and Treatment Protocol for Novel Coronavirus Pneumonia (Trial Version 7). 2020 [cited 2020 May 18th]; Available from: http://www.kankyokansen.org/uploads/uploads/files/jsipc/protocol_V7.pdf.

14. Etienne Campion. Discovery: les experts français qui cherchent un traitement contre le Covid sont-ils sous l’influence des labos ? 2020; Available from: https://www.marianne.net/societe/discovery-les-experts-francais-qui-cherchent-un-traitementcontre-le-covid-sont-ils-sous-l.

15. ROUGUYATA SALL. La chloroquine entre finalement dans l’essai clinique national. 2020; Available from: https://www.mediapart.fr/journal/france/180320/la-chloroquine-entre-finalementdans-l-essai-clinique-national.

16. Branswell, H. WHO to launch multinational trial to jumpstart search for coronavirus drugs. 2020 [cited 2020 May 24th]; Available from: https://www.statnews.com/2020/03/18/who-to-launchmultinational-trial-to-jumpstart-search-for-coronavirus-drugs/.

17. Baker, R. B.C. hospitals taking part in massive WHO COVID-19 treatment study. [cited 2020 May 24th]; Available from: https://www.cbc.ca/news/canada/british-columbia/solidarity-trial-forcovid-19-drugs-1.5528896.

18. Kai Kupferschmidt, J.C. WHO launches global megatrial of the four most promising coronavirus treatments. [cited 2020 May 24th]; Available from: https://www.sciencemag.org/news/2020/03/who-launches-global-megatrial-four-most-promisingcoronavirus-treatments.

19. P., L. CORONAVIRUS: UPDATE ON THE DISCOVERY TRIAL, EUROPE STRUGGLING TO PARTICIPATE. 2020 [cited 2020 May 28th]; Available from: https://www.sortiraparis.com/news/coronavirus/articles/215331-coronavirus-update-on-thediscovery-trial-europe-struggling-to-participate/lang/en.

20. REVORERY: Randomised Evaluation of COVID-19 Therapy. [cited 2020 May 24th]; Available from: https://www.recoverytrial.net/for-site-staff.

21. Wilkinson, E., RECOVERY trial: the UK covid-19 study resetting expectations for clinical trials. BMJ, 2020. 369.

22. Rosenberg, E.S., et al., Association of Treatment With Hydroxychloroquine or Azithromycin With In-Hospital Mortality in Patients With COVID-19 in New York State. JAMA, 2020.

23. Food and Drug Administration (FDA). Remdesivir EUA Letter of Authorization. 1 May 2020; Available from: https://www.fda.gov/media/137564/download.

24. ClinicalTrials.gov. Study to Evaluate the Safety and Antiviral Activity of Remdesivir (GS-5734™) in Participants With Severe Coronavirus Disease (COVID-19). 18 May 2020]; Available from: https://clinicaltrials.gov/ct2/show/NCT04292899?term=Remdesivir&recrs=aef&draw=2&rank=1.

25. ClinicalTrials.gov. Multicenter, Retrospective Study of the Effects of Remdesivir in the Treatment of Severe Covid-19 Infections (REMDECO-19). 18 May 2020]; Available from: https://clinicaltrials.gov/ct2/show/NCT04365725?term=Remdesivir&recrs=aef&draw=2&rank=2.

26. ClinicalTrials.gov. Study to Evaluate the Safety and Antiviral Activity of Remdesivir (GS-5734™) in Participants With Moderate Coronavirus Disease (COVID-19) Compared to Standard of Care Treatment. 18 May 2020]; Available from: https://clinicaltrials.gov/ct2/show/NCT04292730?term=Remdesivir&recrs=aef&draw=2&rank=3.

27. ClinicalTrials.gov. Adaptive COVID-19 Treatment Trial (ACTT). 18 May 2020]; Available from: https://clinicaltrials.gov/ct2/show/NCT04280705?term=Remdesivir&recrs=aef&draw=2&rank=6.

28. ClinicalTrials.gov. The Efficacy of Different Anti-viral Drugs in COVID 19 Infected Patients. 18 May 2020]; Available from: https://clinicaltrials.gov/ct2/show/NCT04321616?term=Remdesivir&recrs=aef&draw=2&rank=7.

29. Goldhill, O. Regulators would consider releasing an unapproved coronavirus vaccine. [cited 2020 May 24th]; Available from: https://qz.com/1852835/fda-would-consider-releasing-anunapproved-coronavirus-vaccine/.

30. Raymond, A.K. The FDA Has Approved a Coronavirus Vaccine for Phase Two Trial. [cited 2020 May 24th]; Available from: https://nymag.com/intelligencer/2020/05/modernas-coronavirusvaccine-approved-for-phase-2-trial.html.

31. Coronavirus vaccine could be ready ‘in a year’ — EU Medicines Agency. [cited 2020 May 24th]; Available from: https://www.dw.com/en/coronavirus-vaccine-could-be-ready-in-a-year-eumedicines-agency/a-53433300.

32. Tidman, Z. Coronavirus vaccine could be ready by this time next year, says EU drugs agency. [cited 2020 May 24th]; Available from: https://www.independent.co.uk/news/health/coronavirusvaccine-cure-treatment-european-medicines-agency-a9513636.html.

